# Implementation of a Novel Remote Physician SBRT Coverage Process during the Coronavirus Pandemic

**DOI:** 10.1101/2020.04.09.20059857

**Authors:** Alex Price, Lauren E. Henke, Borna Maraghechi, Taeho Kim, Matthew B. Spraker, Geoffrey D. Hugo, Clifford G. Robinson, Nels C. Knutson

**Affiliations:** Department of Radiation Oncology, Washington University in St Louis School of Medicine; Department of Engineering Management and Systems Engineering, Missouri University of Science and Technology

## Abstract

**INTRODUCTION:** During the COVID-19 pandemic, alternative methods of care are needed to reduce the relative risk of transmission in departments. Also needed is the ability to provide vital radiation oncological care if radiation oncologists (RO) are reallocated to other departments. We implemented a novel remote RO SBRT coverage practice, requiring it to be reliable, of high audio and visual quality, timely, and the same level of specialty care as our current in-person treatment coverage practice.

**METHODS:** All observed failure modes were recorded during implementation over the first 15 sequential fractions. The time from CBCT to treatment was calculated before and after implementation to determine timeliness of remote coverage. Image quality metrics were calculated between the imaging console screen and the RO’s shared screen. Comfort levels with audio/visual communication as well as overall comfort in comparison to in-person RO coverage was evaluated using Likert scale surveys after treatment.

**RESULTS:** Remote RO SBRT coverage was successfully implemented in 14/15 fractions with 3 observed process failures that were all corrected before treatment. Average times of pre-treatment coverage before and after implementation were 8.74 and 8.51min, respectively. The cross correlation between the imaging console screen and RO’s shared screen was r=0.96 and lag was 0.05s. The average value for all survey questions was above 4.5, approaching in-person RO coverage comfort levels.

**CONCLUSIONS:** Our novel method of remote RO SBRT coverage permits reduced personnel and patient interactions surrounding RT procedures. This may help to reduce transmission of COVID-19 in our department and provides a means for SBRT coverage if ROs are reallocated to other areas of the hospital for COVID-19 support.

## Introduction

The outbreak of the Coronavirus 2019 disease (COVID-19) pandemic has strained the healthcare system in a variety of localities across the globe. In Italy (the European disease epicenter at the time of this writing), intensive care is required for > 15% of infected patients^1^ and the number of hospital beds has been increased by 50% to meet care needs^2^. COVID-19 is highly infectious and a single patient typically infects 2-4 additional persons depending on level of isolation^3,4^. In Italy alone, an additional 20,000 healthcare workers are needed to staff the patient surge, resulting in the shunting of non-ICU and non-internal medicine healthcare providers into the inpatient medicine care space from other specialties^5^. Further, physicians and other providers are at especially high risk of infection--treating physicians comprised 29% off those hospitalized in a study of 138 patients in Wuhan, China--and must be replaced and cared for by their colleagues^6^.

Radiation oncology is not on the front-line of this pandemic but is directly impacted by the disease through anticipatory reductions in staffing to reduce exposure risks, quarantine of infected or exposed providers, and reallocation of the workforce as providers are called upon to reinforce other specialties. Absence of a certain radiation oncology sub-specialist or specialty staff member due to these factors could result in sub-optimal care or lack of availability of certain types of specialty care procedures like stereotactic body radiotherapy (SBRT) ^7,8^. Patients with cancer are also at higher risk of fatality from infection^9,10^, which further underscores the need for increased efforts to minimize transmission within the radiation oncology department to protect these at risk patients. To maximize the availability of staff members, as well as to limit the exposure risks amidst staff and oncology patients incurred through common interactions like in-person SBRT coverage, digital care and coverage techniques must be implemented.

Based on the desired use of remote techniques like telemedicine during the COVID-19 pandemic^11,12^, a novel, digital method to provide physician SBRT coverage for image review was created and implemented in our department. Ease of use and HIPAA compliance were required design features for the deployment of this remote SBRT technology. Equally important was the widespread accessibility of this approach both within our multicenter network and in other clinic systems facing parallel staffing and exposure challenges. Thus, we utilized a commercially available combination of software and hardware to ensure broad reproducibility.

The goal of our remote SBRT coverage technique was to provide reliable, timely, and similar quality specialty care for our patients to reduce the relative risk of infection in our department by encouraging physical distancing amongst personnel and oncology patients. We also aimed to provide a mechanism for our physicians and specialty staff to be allocated elsewhere in the hospital while continuing coverage for radiation oncology care. Here we describe the composition, implementation, and reliability of a digital remote SBRT coverage technique in our radiation oncology department, as well as how it is perceived by our SBRT care team.

## Methods

At pre-COVID-19 baseline, our daily SBRT coverage team consists of two radiation therapists, a medical physicist, and a radiation oncologist. Department policies, developed from national guidelines, require physician presence at every fraction for fusion review, image approval, and to direct treatment, while physics presence is required for all first-fraction treatments to review setup, image registration, motion management, setup corrections, and troubleshooting^13,14^. Apart from these specified roles, both physicians and physicists must also remain available within the department at all times during treatment delivery. Our department delivers an average of 10-15 SBRT treatments per day, the majority of which take place on a specialty stereotactic treatment unit, Edge (Varian Medical Systems, Palo Alto, CA). The typical workflow of coverage is as follows. The patient is set up on the table to marks by the therapy team, and a CBCT is obtained with initial positional match performed by the therapists. The physician is then called and verifies the setup, the shift is sent, and the physician verifies all subsequent images (kV/kV, fluoroscopy, etc) prior to delivery. At first fraction, a physicist is present at time of set up and through delivery. At first (and subsequent) fractions, the physician is called to the machine following initial CBCT match.

To enable transition to remote SBRT coverage in the setting of a global pandemic requiring physical distancing between persons, a commercial grade frame grabber (DVI2USB 3.0, Epiphan Video, Palo Alto, CA) capable of capturing screen resolutions of 1900×1200 at 60fps was connected to the digital video interface (DVI) port on the treatment console through a DVI cord splitter, grabbing the entire display screen from the imaging (treatment console) computer on the Edge. This was then projected into a secondary computer via a USB cable connection between the frame grabber and secondary computer. Using an enterprise installation of Microsoft Teams (Microsoft, Redmond, WA), a video call was placed from the secondary computer and the secondary computer’s screen was shared (displaying the grabbed treatment console screen) with the covering physician in real time. The covering physician can answer the Teams call using either a desktop computer, laptop, tablet, or mobile device. The physician and the treatment machine team (therapists and physicists) must have audio communication to properly communicate; this was fully enabled as long as the device they answer from has audio and microphone capability. To ensure the covering physician has adequate visualization of the treatment images and setup review, the physician can control the secondary computer’s mouse via the Microsoft Teams application. If at any time, an SBRT team member feels uncomfortable with the remote physician process, the physician comes to the machine for an in-person review. The process is illustrated in Figure 1.

**Figure 1.**
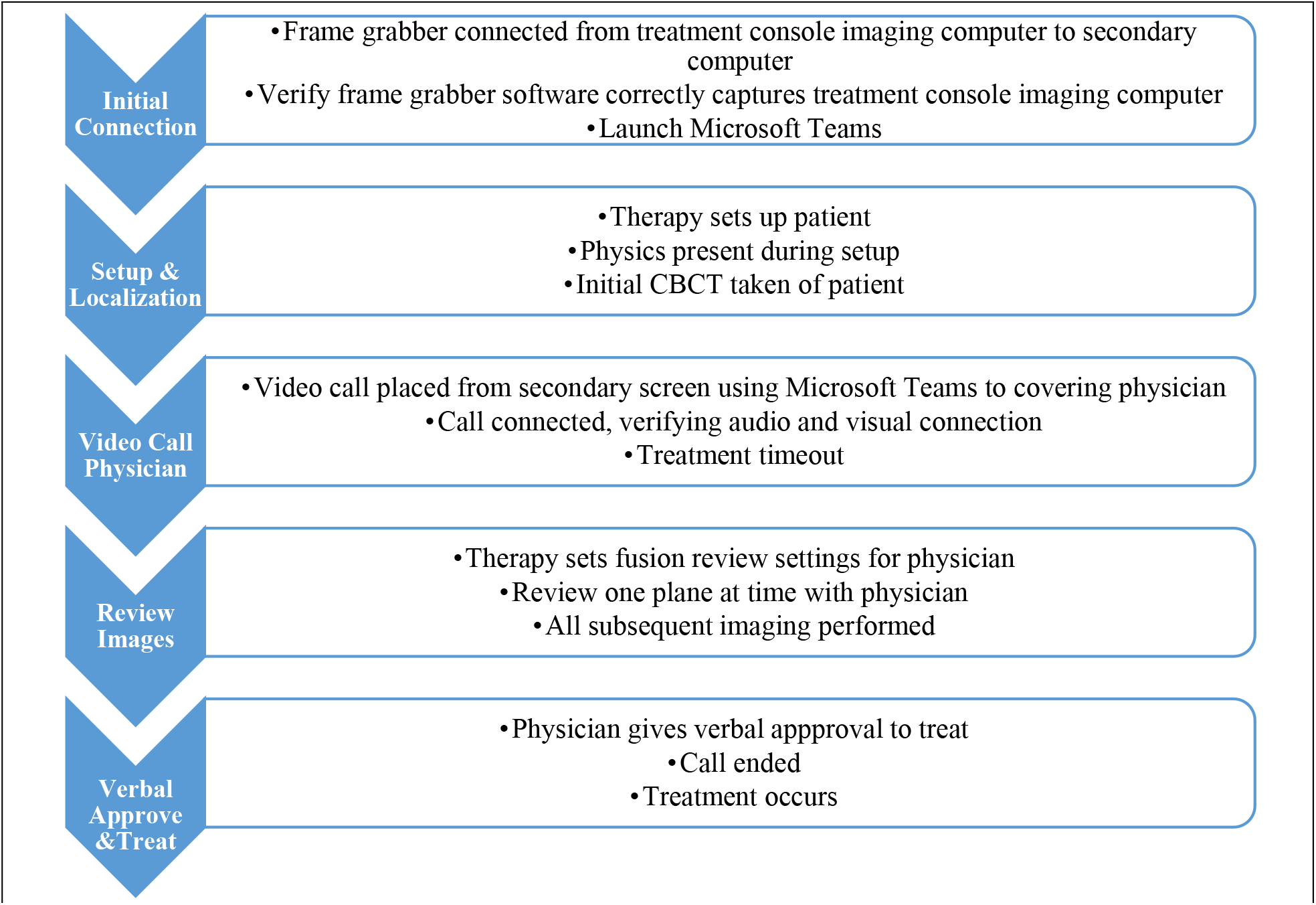
Process map of the remote SBRT coverage workflow.

After implementation of the above process, we sought to evaluate the reliability, timeliness, and quality of our remote SBRT coverage system. Reliability was determined by evaluating the frequency and types of process failures that occurred over the first 15 SBRT fractions completed using remote coverage with the novel system. A list of potential failure modes that were recorded is available in supplementary section A.

Apart from reliability, we also evaluated the timeliness of remote coverage as compared to contemporaneous fractions delivered using our traditional, in-person coverage system. Using the treatment timing stamps from the record and verify system (Aria, Varian, Palo Alto, CA), the time from CBCT acquisition (the time point at which the physician would otherwise be physically called to the machine) to treatment was calculated for 15 fractions after remote SBRT coverage implementation. For comparison, time from CBCT to treatment was also gathered for 15 fractions delivered in the weeks directly preceding remote SBRT coverage. Similar treatment sites were selected for the time analysis before and after remote coverage implementation to limit variations in expected duration. These time stamps represent when the covering physician was contacted and how long they were present at the machine or present on the Teams call.

In addition to reliability and timeliness, we compared both the quantitative and subjective quality of audio and visual (A/V) connection for the remote SBRT coverage platform. Our specific concerns were potential for loss of A/V quality and potential system lag that could degrade coverage quality. Quantitative evaluation of overall A/V quality included comparison of treatment console images versus the physician’s shared screen images using cross-correlation image registration metrics. Image lag was tested similarly, using cross-correlation over time to establish when the frame recorded on the physician’s computer matched the secondary sharing computer’s image frame. The time for the physician’s computer to match the secondary computer was the image lag. With regards to subjective A/V quality, physicians, physicists, and therapists were all surveyed after each SBRT fraction, asking each group to rate A/V quality using a Likert scale (Figure 2).

**Figure 2.**
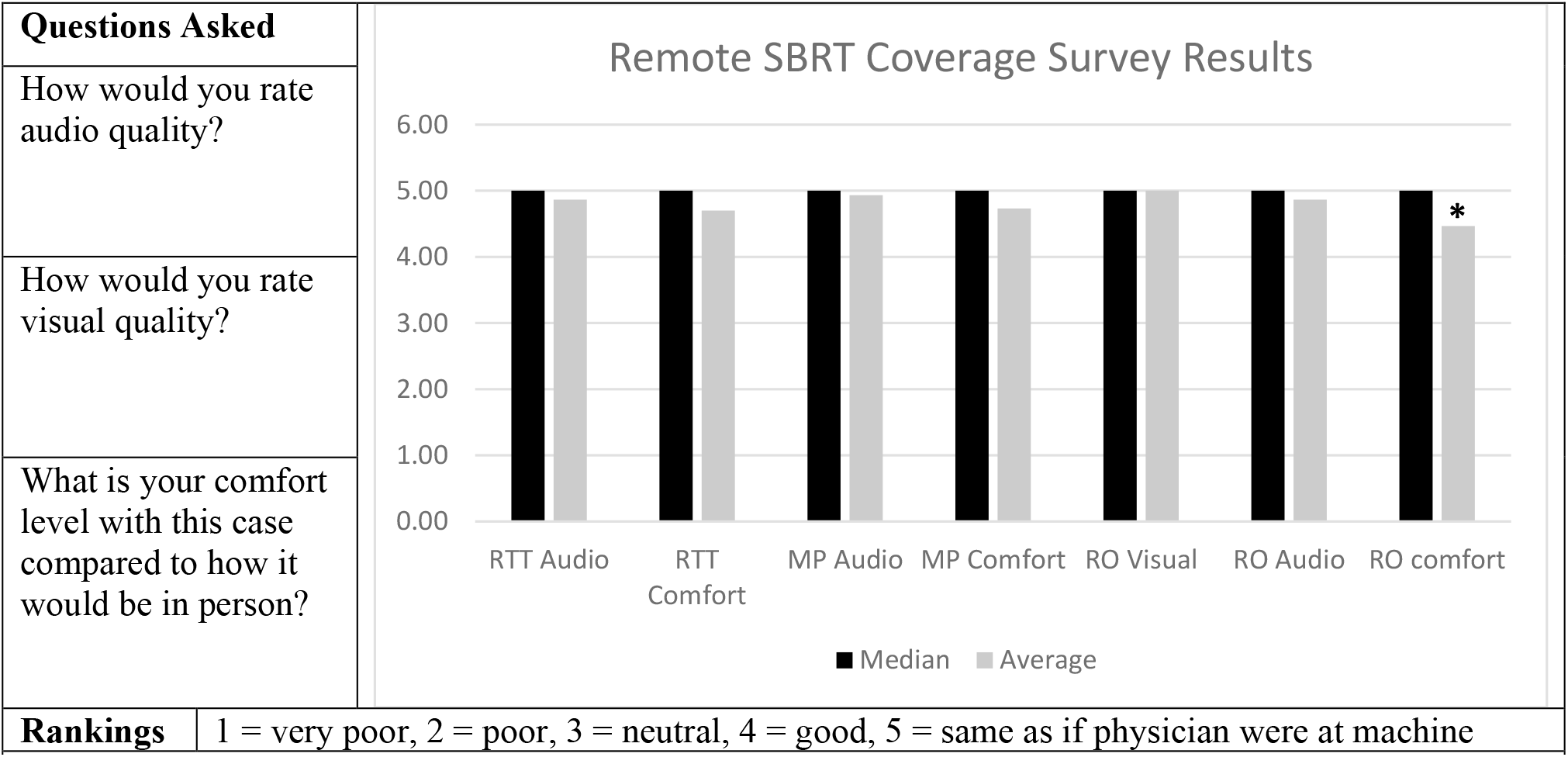
Survey questions and responses. *The responses were statistically different than idealized answers of 5.(p = 0.013)

Finally, we evaluated team member comfort with the novel remote SBRT process. The final question in the post-fraction survey administered to all three personnel groups involved in the SBRT treatment process was “What was your comfort level with this case compared to how it would be in person?” Responses were again recorded using a 5-point Likert scale.

All statistical analyses were completed using SPSS Version 26.0 (IBM Corp., Armonk, NY). For analysis of survey Likert scale data, paired control comparison values of “5” were assumed for all data points, given that surveyed individuals were asked to scale responses such that 1 was “very poor” and 5 was “same as if physician were at the machine”. Unpaired and paired T-tests were utilized for the comparative analyses for the timing data and survey responses, respectively.

## Results

Using a commercially available frame grabber and collaboration platform, we were able to successfully place video calls to the covering physician, sharing the treatment console’s imaging screen, for all 15 SBRT coverage calls. During all 15 remote SBRT coverage calls, a call was never was lost nor were there any noticeable glitches in audio or visual quality. Our physician team was also reliable in terms of responsiveness, never needing to be called more than twice with prompt call-back if missed.

Remote SBRT physician coverage was successfully used for 14 out of the first sequential 15 treatment fractions. We observed occurrence of three system failures from our pre-determined set of potential failure modes during the first 15 cases implementing remote SBRT coverage. The first failure was a near-miss for treatment time out (case description and patient identification), which was not initially performed as planned prior to setup confirmation (preferred timing), but was detected and performed prior to delivery. The second failure was a distracted physician during remote SBRT coverage. This was detected by the treating therapists, who reminded the physician to not multitask during SBRT, with correction prior to delivery. The final failure was a treating physician being uncomfortable with visualization and ability to communicate changes for an abdominal gating case during the confirmation of the gating windows and fiducial fluoroscopy tracking, requiring the physician to come to the machine to clarify setup. This resulted in a physician comfort level of 2, or poor.

The disease sites treated for the reported first 15 remote coverage fractions were abdomen (n=4), bony extremity (n=4), brain (n=3), spine (n=2), lung (n=1), and neck (n=1). The comparison cases selected from the preceding weesk using in-person physician coverage were intentionally selected to be of an identical disease site distribution.

The average time from CBCT to the time of treatment for the 15 fractions before implementation was 8.74min (median 7.93min). The average time from CBCT to the time of treatment for the 15 fractions after implementation was 8.51min (median 7.48min). These were not statistically different (p = 0.504). A distribution of response times are shown in Figure 3. The two cases requiring the most time after remote treatment implementation were the abdominal gating case, where the physician ultimately came to the machine, and an additional abdominal case where confirmation of setup required comparison to the simulation 4DCT image set. When excluding these two longest remote coverage cases (Figure 3), the average time for remote coverage was reduced to 7.28 min (p = 0.088 for comparison to in-person coverage). The median amount of time that physicians were on the remote coverage Teams call was 4.43min.

**Figure 3.**
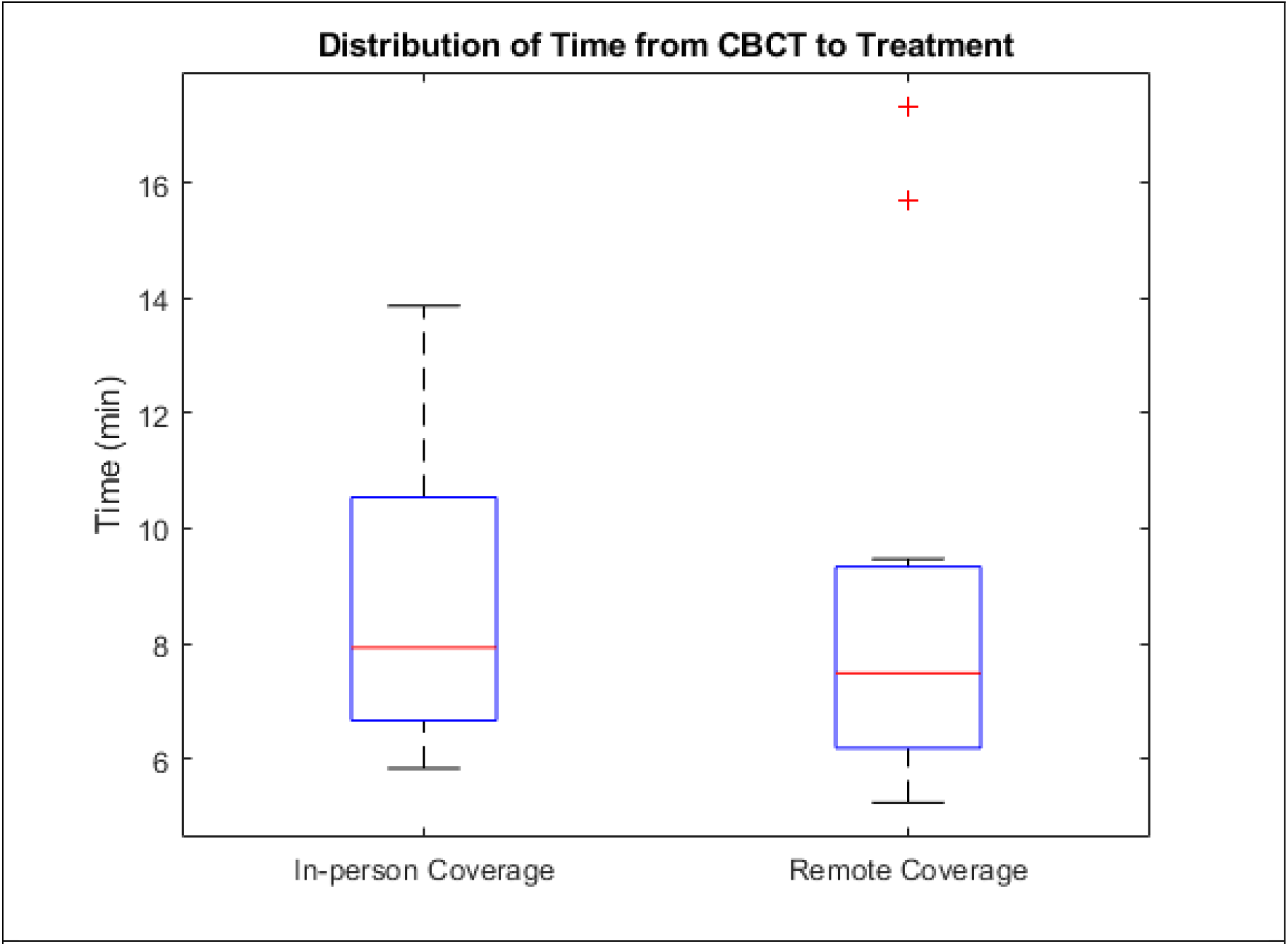
Physician coverage times before remote SBRT coverage (left) and after remote SBRT coverage implementation (right). The red ‘+’ are statistical outliers.

In terms of quantitative image quality, the cross correlation between the treatment console’s imaging screen and the shared frame grabbed screen via Microsoft Teams was R = 0.96. The lag in Teams video sharing was 0.05s (50ms). An example screenshot from a physician’s mobile device (iPhone X) during remote coverage is shown in Figure 4. Two-finger pinch zoom-in and zoom-out was also feasible on touch-screen devices, without perceptible lag. A demonstration of the A/V quality and the workflow can be viewed in supplementary section B.

**Figure 4.**
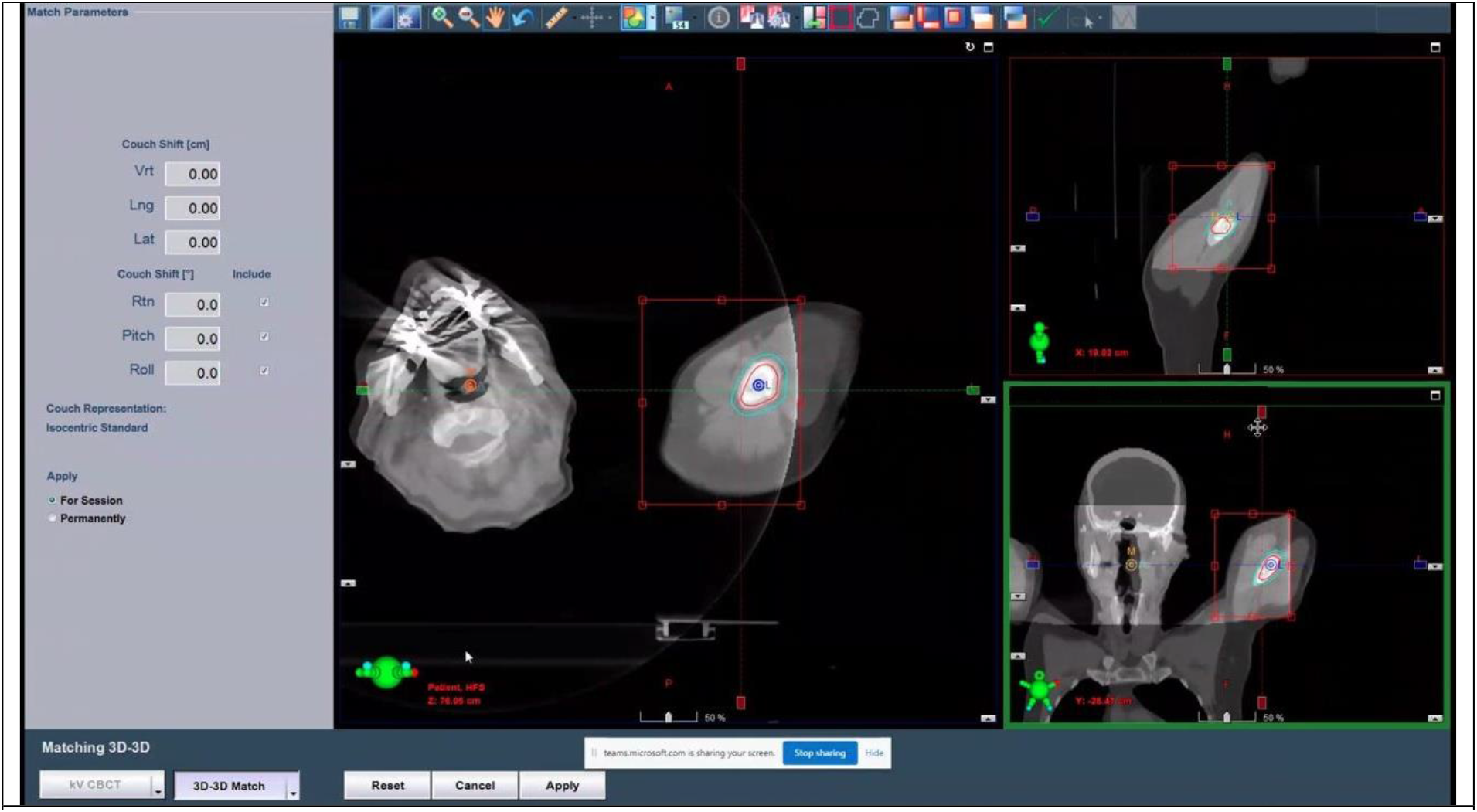
Example of covering physician’s view on mobile device (iPhone X).

The remote SBRT survey questions and responses for A/V quality and level of comfort are shown in Figure 2. The only category that was statistically significantly different from in-person coverage was physician comfort (Figure 2, p = 0.013). The lowest ranked comfort level was 2, which occurred once. There were 5 responses with scores of 4, and the majority of these 4s were because the physician would have ideally changed a windowing level or fusion review visualization setting (i.e. floating vs. gray/white overlay vs. checker-box). Nevertheless, the median response was 5 across all categories, including for audio quality.

## Discussion

We have successfully implemented remote SBRT physician coverage within our department for a variety of treatment sites. The tools that we use are available to most radiation oncology departments and can be implemented quickly with a few wire connections and installations of commercial applications. Remote SBRT coverage will allow for flexibility in response as staffing resources are shunted or reallocated during the COVID-19 crisis, while providing similar quality to in-person SBRT coverage. All members of the treatment team received the process positively and were invested in making improvements as we continued to refine and gain experience with the process.

Remote SBRT coverage is feasible with minimal observed potential failure modes based on our initial implementation data. Of the first 15 remote coverage cases attempted, only 1 case ultimately required physician presence. Specifically, nuances of communication using remote instructions for gating windows for a gated abdominal case led to physician discomfort with remote coverage, and the physician instead opted to be physically present at the console for more direct communication. Other observed failure modes were a distracted physician (multiple phone messages while covering via Teams mobile app call) and a nearly-missed pre-treatment time-out. Both of these were detected and corrected prior to treatment. Formal FMEA analysis is the optimal approach prior to implementation of novel technologies in a medical environment^15^. However, given the urgency of creating a remote coverage process in the pandemic setting, this will instead be formally carried out after remote SBRT implementation.

With regards to timeliness, remote SBRT coverage compared favorably to our in-person process. Time delays are critical to avoid for successful deployment of the tool, as this would prolong patient time on-table, which can result in loss of setup accuracy^16^. We found that the time of CBCT to the time of treatment was not statistically different for remote coverage compared to in-person coverage. It is important to note that during this short time period of investigation most of the cases analyzed were not our institution’s typical SBRT disease sites (majority lung and brain), lending to the possibility that pre-treatment coverage could be faster for standard SBRT disease sites compared to our current process of requiring the physician to be physically present. Importantly, remote coverage physically removed physicians from the machine, which would otherwise be an average of 4.5min for in-person coverage at each case. This may reduce the relative risk of infectious exposure across personnel and patients. Physicians were able to remote review from areas in the department such as nursing wings, their office, and outside a consult room. We also found it helpful to inform the covering physician when the treatment team was bringing the patient into the room, allowing the physician to expect a call and increase the answer rate.

Our image quality degradation through Microsoft Teams via the frame grabber was minimal and maintained high correlation within the hospital network, meaning the images were almost identical. The lag was 50msec, which is in the range of what is perceived as “instantaneous action” to a user based on published literature^17,18^. Our image quality metrics as determined by post-treatment surveys demonstrate non-inferiority to the physician’s visual perception if he or she were present at the machine.

The comfort level amongst all members of the treatment team approached the in-person SBRT coverage comfort levels, demonstrating that our SBRT team felt comfortable proceeding with treatment despite not having a physician physically present. The lowest comfort level reported for any fraction was a 2, due to inconsistencies between the CBCT and fluoroscopic gated images. Despite a low comfort level, we were able to correctly identify that the physician was needed at the machine and had minimal wait time for the physician to be present (approximately a minute or less). In the future, we look to identify cases where remote SBRT coverage may be challenging, such as first fraction abdominal gating cases or for lesions that have high potential for poor detectability even if in person (e.g. peridiaphragmatic lung lesions). Additional time-outs during this process are necessary to ensure physicians have all needed information. Ordinarily, time out is performed before the patient is placed on the table and again when physician arrives for coverage. With in-person coverage, the physician has the opportunity to pull up patient charts and supplementary imaging alongside the treatment console on a separate computer from the Microsoft Teams computer. In the remote setting, especially by mobile app, a second time-out to transmit this information about the case to the physician is critical. Having available resources at the ready, including prior fraction notes, planning scans, etc. to answer whatever questions that the physician has may be helpful. Additional challenges include the greater importance of using direct verbal communication, as common non-verbal cues are lost with remote coverage. Limiting distractions from competing physician tasks is also critical to ensure quality and future steps may include a formal coverage script where physicians maintain an active verbal role in the process to ensure continued attentiveness.

Future steps in our process include performing a formal FMEA analysis of remote SBRT coverage for our physicians. We would also like to perform a study investigating remote coverage differences for a larger set of patients over a longer period of time. This work allows us to explore avenues of real-time training and recording across our multi-institutional sites during deployment of new techniques for debrief and feeback. It also provides a platform for real-time collaboration across multiple individuals with expertise in a variety disease sites despite not being physically available at the time of treatment. At this current stage, we were able to quickly implement this novel technology, performing a brief yet informative and important study in the management of COVID-19 for the healthcare system.

## Conclusion

In our study, we were able to provide remote SBRT coverage that is of high visual and audio quality, reliable, timely, and similar to in-person physician SBRT coverage. Because of this, we may reduce the relative risk of transmitting COVID-19 amongst our colleagues and patients. Our physicians were able to perform remote SBRT coverage in various locations in our department and hospital, meaning coverage is possible even if physicians are allocated to other areas in the hospital. Remote coverage permits continued high-quality care for our oncology patients, maximizing physicians as resources for other types of patient care during the COVID-19 crisis, and minimizes exposure risk to our staff members and patients.

## Data Availability

Data is not publicly available.

## Statement on Ethics Board Approval

This research was approved by our institutional review board. The IRB ID# is 202004015.

